# Viral dynamics of Omicron and Delta SARS-CoV-2 variants with implications for timing of release from isolation: a longitudinal cohort study

**DOI:** 10.1101/2022.04.04.22273429

**Authors:** Tara C. Bouton, Joseph Atarere, Jacquelyn Turcinovic, Scott Seitz, Cole Sher-Jan, Madison Gilbert, Laura White, Zhenwei Zhou, Mohammad M. Hossain, Victoria Overbeck, Lynn Doucette-Stamm, Judy Platt, Hannah E. Landsberg, Davidson H. Hamer, Catherine Klapperich, Karen R. Jacobson, John H. Connor

**Author notes:** **Contact Information – Corresponding and Alternate Corresponding Authors:** Tara C. Bouton, Boston University School of Medicine, Section of Infectious Diseases, 801 Massachusetts Ave, 2^nd^ Floor, Boston, MA 02119, John Connor, National Emerging Infectious Diseases Laboratories, 620 Albany Street, Boston, MA 02118. Equal manuscript contributions (Catherine Klapperich, Karen R. Jacobson, John H. Connor).

## Abstract

**Background:** In January 2022, United States guidelines shifted to recommend isolation for 5 days from symptom onset, followed by 5 days of mask wearing. However, viral dynamics and variant and vaccination impact on culture conversion are largely unknown.

**Methods:** We conducted a longitudinal study on a university campus, collecting daily anterior nasal swabs for at least 10 days for RT-PCR and culture, with antigen rapid diagnostic testing (RDT) on a subset. We compared culture positivity beyond day 5, time to culture conversion, and cycle threshold trend when calculated from diagnostic test, from symptom onset, by SARS-CoV-2 variant, and by vaccination status. We evaluated sensitivity and specificity of RDT on days 4-6 compared to culture.

**Results:** Among 92 SARS-CoV-2 RT-PCR positive participants, all completed the initial vaccine series, 17 (18.5%) were infected with Delta and 75 (81.5%) with Omicron. Seventeen percent of participants had positive cultures beyond day 5 from symptom onset with the latest on day 12. There was no difference in time to culture conversion by variant or vaccination status. For the 14 sub-study participants, sensitivity and specificity of RDT were 100% and 86% respectively.

**Conclusions:** The majority of our Delta- and Omicron-infected cohort culture-converted by day 6, with no further impact of booster vaccination on sterilization or cycle threshold decay. We found that rapid antigen testing may provide reassurance of lack of infectiousness, though masking for a full 10 days is necessary to prevent transmission from the 17% of individuals who remain culture positive after isolation.

**Main Point:** Beyond day 5, 17% of our Delta and Omicron-infected cohort were culture positive. We saw no significant impact of booster vaccination on within-host Omicron viral dynamics. Additionally, we found that rapid antigen testing may provide reassurance of lack of infectiousness.

## Background

Individuals with SARS-CoV-2 infection have been told to self-isolate to avoid transmission to others. Early pandemic United States Centers for Disease Control and Prevention (CDC) guidelines for isolation were based on estimates of the duration of infectivity, with early studies showing rare culture positivity and transmission beyond 10 days in immunocompetent hosts.^1^ In January 2022, US CDC guidelines shifted to recommend 5 days of strict isolation from symptom onset or positive test if asymptomatic, followed by an additional 5 days of strict mask wearing.^2^ This newer guidance was based on SARS-CoV-2 transmission studies showing that most transmission occurs early in the course of infection,^3,4^ the increased awareness of the mental health, economic, and social impacts of prolonged isolation,^5^ and that only 25-30% of cases truly isolate for a full 10 days.^6^

A proposed strategy to limit time in strict isolation of SARS-CoV-2 infected individuals has been to use antigen rapid diagnostic testing (RDT) as a proxy for infectiousness.^2^ A test to return program in a Massachusetts school district found positivity rate of 35% when performed day 6 from symptom onset or positive test, concluding that such a program helps identify those safest to return to the classroom prior to 10 days.^7^ Modeling efforts have further suggested that antigen RDT could be used to both reduce the self-solation period while preventing ongoing disease transmission.^8^

New SARS-CoV-2 variants have shown increased transmissibility, potentially impacting best practices for isolation guidelines. Notably, the Omicron variant may be up to three times more transmissible than the Delta variant,^9^ with a shorter incubation period,^10^ but lower viral loads at diagnosis^11^ than previously circulating SARS-CoV-2 lineages. Early SARS-CoV-2 viral challenge data with wild-type virus in seronegative, healthy volunteers showed on average nasal culture clearance by day 7 with culture positivity up to 12 days from SARS-CoV-2 exposure.^12^ How long vaccinated individuals, both primary series and boosted, can transmit virus and kinetic difference by variant remain unclear.

We aimed to document within-host viral dynamics of two recent SARS-CoV-2 variants, Delta and Omicron, during the period when most individuals would leave isolation based on CDC guidelines. We recruited participants from a university campus which had a multi-faceted surveillance testing and COVID-19 control program^13^. We compared infection with Omicron versus Delta, the vaccination and booster status of the infected individual, and the difference from test date and symptom onset date on detectable virus and culture positivity.

## Methods

The SARS-<u>Co</u>V-2 <u>Vi</u>ral <u>D</u>ynamics <u>Post-vaccination</u> Study (CoViD Post-vax) is an observational cohort study that, beginning in November 2021, has been enrolling Boston University (BU) students, faculty, and staff. Participants are enrolled after a verbal assent or electronic self-consent process following diagnosis with SARS-CoV-2 by PCR as a part of the BU SARS-CoV-2 screening program which includes regular testing 1-2 times per week. Participants undergo an initial clinical questionnaire and thereafter self-performed daily symptom screens and anterior nasal swabs with electronic medical record and diagnostic specimen laboratory data abstraction performed by study staff.

Following study enrollment, participants can also opt into an antigen RDT sub-study. For those who provide verbal assent, they are provided with three Abbott BinaxNOW kits to self-perform according to manufacturer directions on days 4-6 from diagnosis. Participants are asked to upload a photograph of the test strip at the time it is read into our REDCap database (version 12.1.1, Vanderbilt University), where study staff then interpret the results.

Diagnostic isolates are tested with RT-qPCR using a two-target SARS-CoV-2 assay with RNase P control^1^ as a part of the BU SARS-CoV-2 surveillance program at the BU Clinical Testing Laboratory.^3^ Positive samples are then transferred to the National Emerging Infectious Diseases Laboratories (NEIDL), where viral genomes are sequenced using a modified ARTIC primer–based protocol for amplification and the Illumina platform for sequencing.^14^ For participants whom diagnostic isolate sequencing was not yet available (N=32), variant determination was made based on the timing of the diagnostic isolate: all infections after December 28, 2021 were considered Omicron because BU sequencing surveillance activities detected <1% Delta circulating.^11^ Study isolates are collected in viral transport media and frozen following collection. Samples are then analyzed in two different ways. First, the presence of SARS-CoV-2 viral nucleic acid in the sample is determined by SARS-CoV-2 CDC N1 RT-PCR and second, 200 microliters of sample are incubated with Caco2 cells for 4 days. At the end of culturing, cells are fixed, and the presence or absence of SARS-CoV-2 viral replication is determined through indirect immunofluorescence microscopy using an antibody recognizing SARS-CoV-2 N protein. Cultures are determined to be positive for viral growth if they show cytoplasm-restricted fluorescence signal above background.

In this analysis, we compared demographic and clinical characteristics of participants infected with the Delta and Omicron SARS-CoV-2 variants. Fully vaccinated individuals had completed an initial vaccine series with either a single vector or two mRNA vaccines listed for emergency use by the World Health Organization (WHO), with the exception of one participant who received a non-WHO listed vaccine and was subsequently boosted with an mRNA vaccine. We compared culture positivity beyond day 5, time to culture conversion, and duration of positive cultures when calculated from date of diagnostic test to symptom onset. Culture conversion was defined as day of first negative culture result with no subsequent positive cultures. For the purposes of all cycle threshold (Ct) comparisons, we used the N1 genes from both assays. We plotted scatter plots and fit mixed-effect models to assess the relationship between Ct values and time since diagnosis and symptom onset, and by variant and vaccination status. Curve fits were performed using generalized additive mixed models with random intercept and a cubic B-spline with four knots, to incorporate individual participants’ Ct trajectories. To look at time to culture conversion for the whole cohort and by variant and vaccination status, we generated Kaplan Meier plots where the event was defined as the first negative SARS-CoV-2 culture with no subsequent positives. Heat-treated baseline diagnostic isolates were not culturable and were presumed positive for the purposes of this analysis. When the first recorded negative culture was preceded by missed tests, we used interval censoring with the earliest point of the interval being the time of the last positive test or diagnosis if there were no positive cultures recorded. Additionally, we compared median Ct values at diagnosis by variant and vaccine status, using Wilcox non-parametric tests. Finally, for the antigen RDT sub-study, we calculated the sensitivity and specificity of the Abbott BinaxNow compared to culture positivity at days 4 through 6 from diagnosis and plotted Ct values by day from diagnosis and RDT result. Analyses were conducted with R (version 4.0).

The study was approved by both the BU Charles River Campus Institutional Review Board and the BU Medical Campus Institutional Review Board.

## Results

Of 92 SARS-CoV-2 RT-PCR positive participants enrolled in the study cohort, 17 (18.5%) were infected with the SARS-CoV-2 Delta variant and 75 (81.5%) with Omicron (Table 1). Most participants (N=65, 69.1%) had symptoms at the time of diagnosis. While a greater proportion individuals infected with Omicron had received a booster vaccine compared to those infected with Delta [42.7% (N=32) vs 5.9% (N=1), p=0.004, Table 1), all participants had completed the initial SARS-CoV-2 vaccine series (Table 1).

**Table 1:** Characteristics of participants overall and by SARS-CoV-2 variant

|  | Total (N=92) | Delta (N= 17) | Omicron (N= 75) | p-value |
| --- | --- | --- | --- | --- |
| Age |  |  |  | 0.485 |
| Mean (SD) | 22 (3) | 22 (2) | 22 (3) |  |
| Range | 18 - 33 | 19 - 25 | 18 - 33 |  |
| Sex, N (%) |  |  |  | 0.396 |
| Male | 35 (38.0) | 8 (47.1) | 27 (36.0) |  |
| Female | 57 (62.0) | 9 (52.9) | 48 (64.0) |  |
| Race, N (%) |  |  |  | 0.730 |
| White | 51 (56.0) | 9 (52.9) | 42 (56.8) |  |
| Black | 6 (6.6) | 2 (11.8) | 4 (5.4) |  |
| Asian | 29 (31.9) | 5 (29.4) | 24 (32.4) |  |
| Multiracial | 5 (5.5) | 1 (5.9) | 4 (5.4) |  |
| Missing | 1 | - | 1 |  |
| Vaccination Status,<br>N (%) |  |  |  | 0.004 |
| Fully Vaccinated,<br>not boosted | 59 (64.1) | 16 (94.1) | 43 (57.3) |  |
| Fully vaccinated,<br>boosted | 33 (35.9) | 1 (5.9) | 32 (42.7) |  |
| Vaccine Type, N (%) |  |  |  | 0.196 |
| Pfizer | 61 (66.3) | 10 (58.8) | 51 (68.0) |  |
| Moderna | 22 (23.9) | 4 (23.5) | 18 (24.0) |  |
| Janssen | 3 (3.3) | 2 (11.8) | 1 (1.3) |  |
| Other<sup>a</sup> | 6 (6.5) | 1 (5.9) | 5 (6.7) |  |
| Symptomatic at baseline, N (%) |  |  |  | 0.241 |
| No | 27 (30.9) | 3 (17.6) | 10 (30.9) |  |
| Yes | 65 (69.1) | 14 (82.4) | 65 (69.1) |  |

<sup>a</sup>Covishield (Serum Institute of India), AstraZeneca (Oxford), Sinovac (CoronaVac), Sinopharm
BIBPVeroCell

Overall, 84% of participants from diagnostic test and 71% of participants from symptom onset had culture converted (no growth) by day 6 (Table 2). Overall, 48% (N=44) of participants never had a culture positive research isolate. We next considered the impact of calculating the isolation period as days from diagnosis (common in large institutional settings, like universities) in comparison to days from symptom onset (US CDC guidance)^2^ on viral dynamics at release from isolation. In this cohort, almost half of the participants were diagnosed after symptom onset (N=38, 41.3%), while ten (10.9%) tested positive while pre-symptomatic. Using time from symptom onset rather than time from diagnosis shifted the axis right for those diagnosed after symptom onset and left for pre-symptomatic participants. In comparison to within-host viral load decay over time from diagnosis (Figure 1a), the mixed-effect model of within-host viral load decay from symptom onset (Figure 1b) shows a plateau in the pre-symptomatic period (days - 3 to 0) followed by a more gradual viral decline. Similarly, adjusting the viral culture results from days since diagnosis (Figure 1c) to days from symptom onset (Figure 1d) reveals a considerable drop in culture positivity following day 4 with associated reduction in the maximum duration of culture positivity by 3 days (from 15 days to 12 days, Table 2).

**Table 2:** Culture positivity when calculated from day of diagnosis in comparison to day of symptom onset

|  | Time From Diagnosis<br>(N=92) | Time From Symptom<br>Onset (N=92) | p-value |
| --- | --- | --- | --- |
| Individuals with a culturable isolate >5 days |  |  |  |
| Culture converted by day 6, N (%) | 77 (84) | 65 (71) | 0.05 |
| Culture positive >5 days, N (%) | 10 (11) | 16 (17) | 0.38 |
| Missing, N (%) | 5 (5) | 11 (12) | 0.19 |
| Last culturable day, days | 15 | 12 | NA |

**Figure 1:**
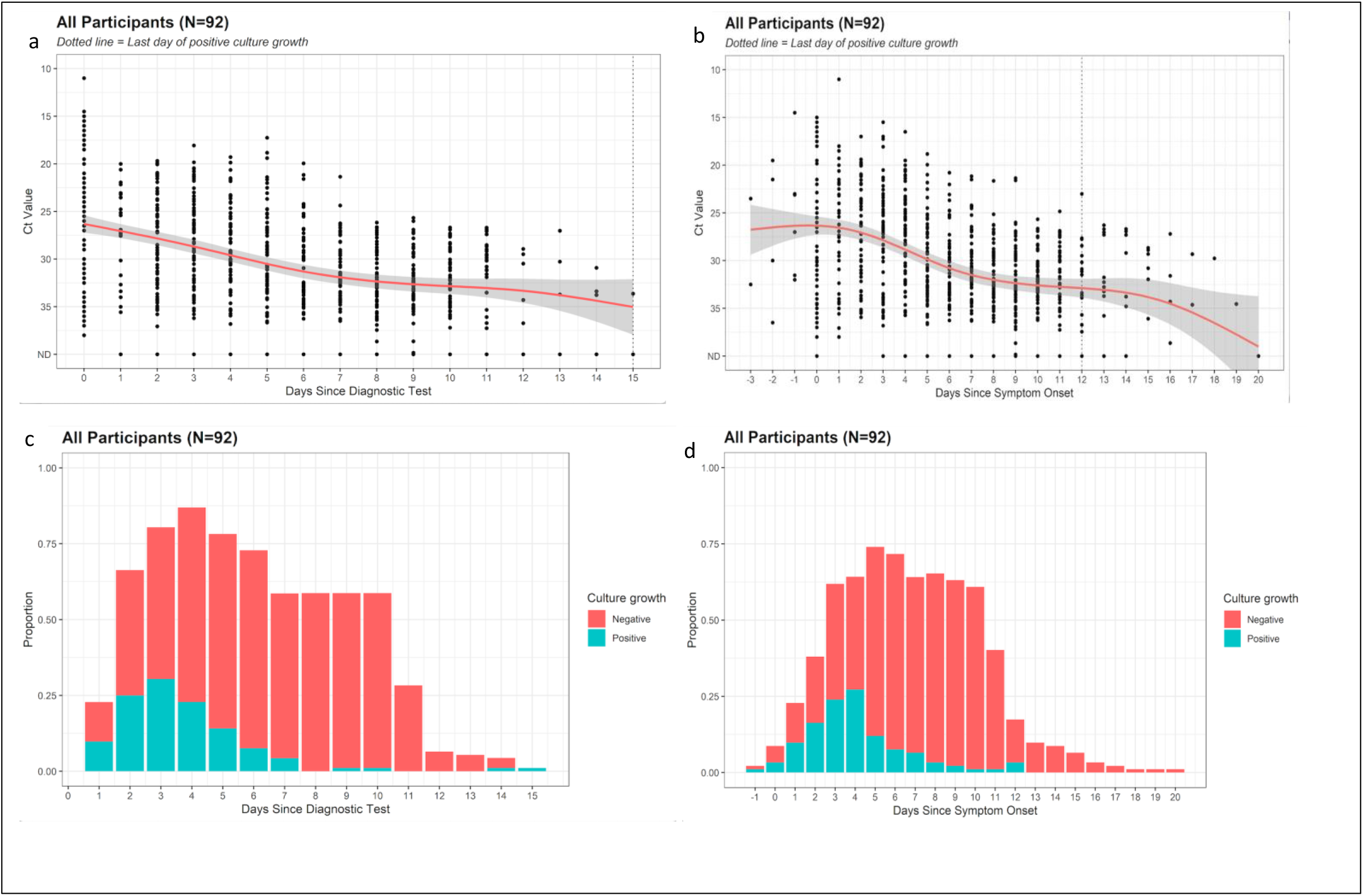
Progression of polymerase chain reaction N1 cycle threshold values from (a) diagnosis and (b) symptom onset, and culture growth from (c) diagnosis and (d) symptom onset.

There were no significant differences in time to viral culture conversion between Delta and Omicron variants, even when stratified based on whether participants had received a COVID-19 booster vaccination or had completed the initial vaccine series alone (Figure 2B). The trends for within-host viral load decay over time were similar in those who completed the initial vaccine series and were infected with the Delta variant (Figure 3B) and the Omicron variant (Figure 3C). However, for participants who had received a COVID-19 booster vaccination, there was a trend towards slower with-in host viral load decay (Figure 3D), though daily culture conversion rates were similar (Figure 2B) and RT-PCR Ct was higher in those boosted with Omicron at diagnosis (median 27.5) than in those fully vaccinated with either Delta (median 18.8, p=0.004) or Omicron infections (median 26.4, p=0.36, Supplementary Figure 1).

**Figure 2.**
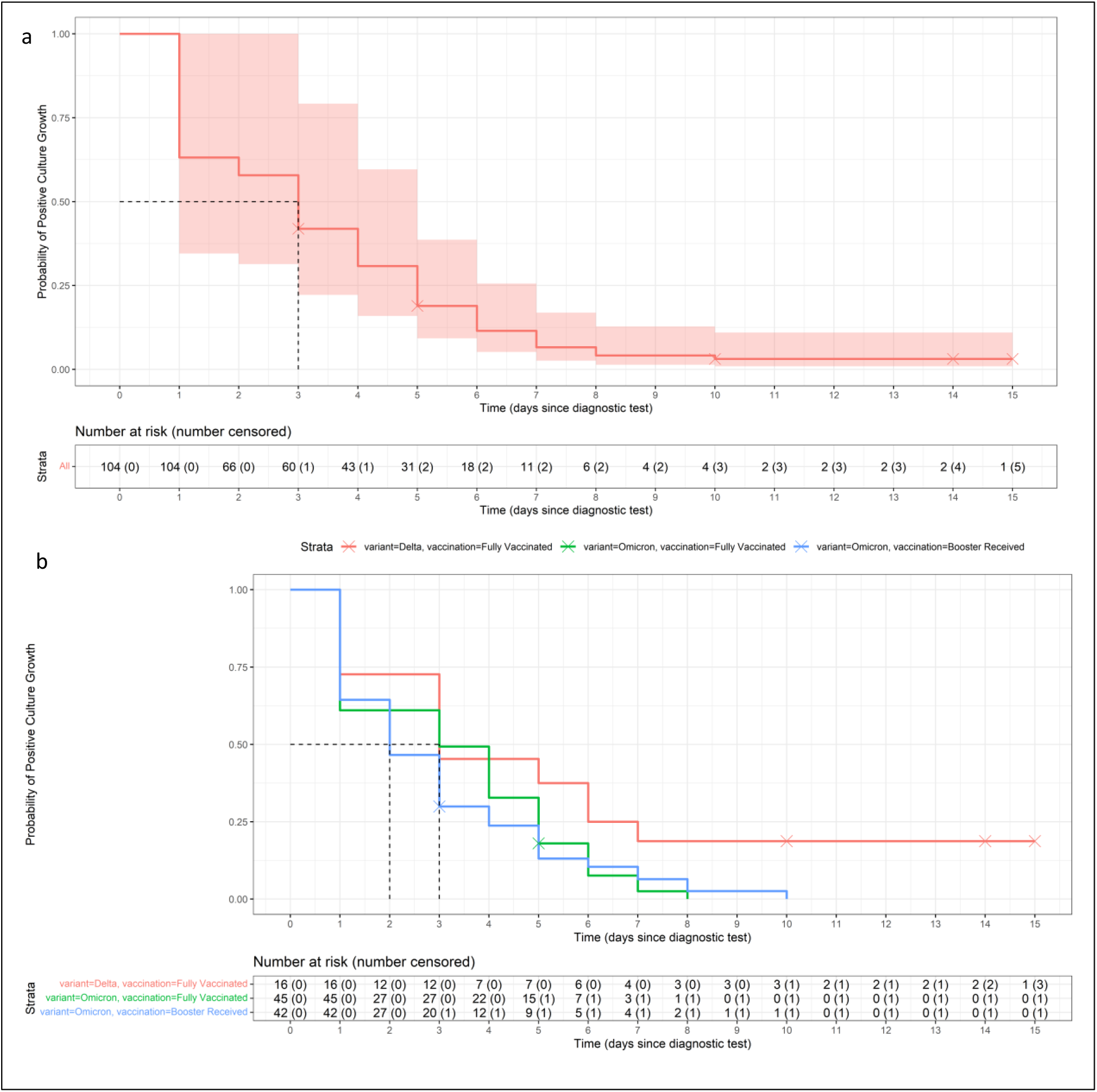
Kaplan-Meier curves indicating days from diagnosis to negative viral culture for all participants (a) and by SARS-CoV-2 variant and vaccination status (b).

**Figure 3:**
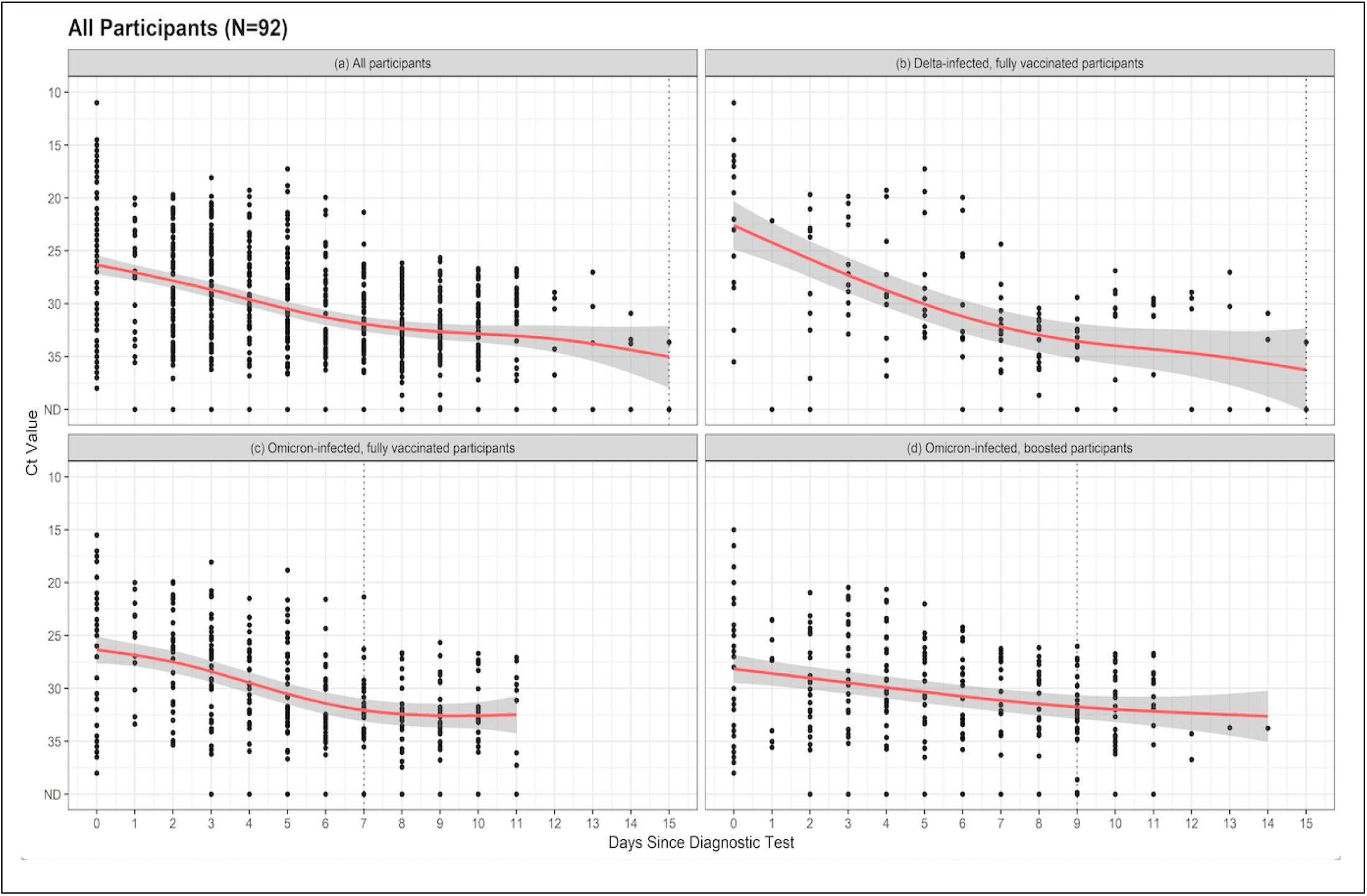
Polymerase chain reaction N1 cycle threshold at days since symptom onset for (a) all participants, (b) delta-infected, fully vaccinated participants, (c) Omicron-infected, fully vaccinated participants, and (d) Omicron-infected, boosted participants.

For the sub-set of 14 participants in the RDT sub-study, the sensitivity and specificity of the Abbott BinaxNOW test were 100% and 86% respectively at day 4 through 6 from diagnosis when compared to SARS-CoV-2 culture (N=32 isolates, Table 3), yielding a negative predictive value of 100% and positive predictive value of 50%. RT-PCR Ct values were higher in those who tested negative on the RDT (Supplementary Figure 2).

**Table 3:** Sensitivity and specificity of rapid antigen testing when compared to culture growth at days 4 through 6 from SARS-CoV-2 diagnosis.

| Days since |  |  |  |  |  |  |  |
| --- | --- | --- | --- | --- | --- | --- | --- |
| Diagnostic |  | True | True | False | False |  |  |
| Test | N | Positive | Negative | Positive | Negative | Sensitivity | Specificity |
| Day 4 | 10 | 2 | 6 | 2 | 0 | 1 | 0.75 |
| Day 5 | 12 | 2 | 9 | 1 | 0 | 1 | 0.9 |
| Day 6 | 10 | 0 | 9 | 1 | 0 | NA | 0.9 |
| Overall | 32 | 4 | 24 | 4 | 0 | 1 | 0.86 |

## Discussion

Our findings suggest that in young, healthy, vaccinated adults, the majority have a limited infectious period for SARS-CoV-2 based on culturable virus and rapid viral decay. We found that beyond 5 days from symptom onset only 17% remained culture positive. Our median time to Omicron culture conversion was 2 days (IQR: 1-5) for boosted participants with Omicron, 3 days (IQR: 1-5) for vaccinated, unboosted participants with Omicron, and 3 days (IQR: 1-6.5) for participants with Delta. This is notably earlier than the median 6 days to culture conversion reported in an older cohort with more medical comorbidities.^14^ Our results are similar to data from the National Basketball Association cohort which found 40% with cycle thresholds of less than 30 at day 5, though this study did not examine culture positivity.^15^

Whether we calculated days to culture clearance from date of diagnosis or from symptom onset led to differences on likelihood that individuals released from isolation were still infectious and how quickly some were released. Reliance on time from diagnosis for determining duration of isolation kept 41% of people in isolation longer than necessary (using culture conversion as a measure of non-infectiousness). Additionally, for the 10 (11%) who were pre-symptomatic at the time of diagnosis, these individuals were released from strict isolation before they had reached the CDC-recommended 5-day point, although only two (20%) remained culture positive at that point which was similar to the overall cohort fraction. It is much easier programmatically for large institutions with serial testing programs to rely on time from diagnosis, a number they capture through these programs, rather than symptom development, which requires additional follow up for those without symptoms and honest reporting of those diagnosed. It is reassuring to see that only 17% of our cohort failed to culture convert by day 6, however it will be key to continue to reinforce strict masking for the full 10-days from symptom onset, especially in scenarios with asymptomatic screening strategies.

We found no major differences in culture conversion or viral load decay between delta and omicron variants. This is consistent with previous data showing no major differences in Omicron infection duration when compared to Delta^16^ or time to culture or PCR conversion between the two variants.^15^ Similar to the findings of Boucau et al.^15^, we found no significant difference in culture or PCR conversion in boosted participants when compared to those who were fully vaccinated. This suggests that the population level protection provided by COVID-19 booster vaccination doses against Omicron^17,18^ result from protection against infection rather than altered viral kinetics in vaccinated individuals causing reduced transmission or more rapid clearance.

Among participants who used an antigen RDT in days 5-7 following SARS-CoV-2 diagnosis, RDT had perfect negative predictive value and sensitivity when compared to culture. While we used a single type of antigen RDT in our sub-study, the Abbott BinaxNOW has been shown to successfully detect the Omicron variant.^19^ Our findings are similar to early reports in other settings suggesting 30-55% positive antigen RDT between days 5-10 of illness.^20,21,7^ One study, using a different antigen test, showed higher antigen RDT positivity (75%), but similar culture positivity (35%) at day 6.^20^ Given the similar negative predictive value, the work by Cosimi et al.^22^ highlights possible differing performance characteristics at the end of infection across tests. Our preliminary findings suggest that a young, healthy, fully vaccinated individual can be reassured with negative antigen RDT that they are unlikely to be shedding viable virus towards the end of their infection. However, reliance on antigen RDT for release from isolation would also result in potential prolongation of isolation for those who do not have culturable virus.

Our study is limited in its generalizability, as most participants were young, healthy, and all had completed at least the initial COVID-19 vaccination series. Additionally, many participants had recently received booster vaccination when omicron began to circulate possibly resulting in higher neutralizing antibody titers than those further out from their last vaccine. Additionally, our work was likely not well powered to determine differences between Delta and Omicron variants stratified on COVID-19 vaccination status. While our study is strengthened by our daily sampling and culture data, it is not clear how well culture positivity in the laboratory correlates to transmissibility of SARS-CoV-2 at the end of an infection. More work needs to be done to understand why certain immunocompetent individuals have prolonged culture positivity and how to predict which cases will remain at risk of transmitting beyond the isolation period. Regardless, it is clear that even in a young and healthy cohort, with presumably optimal response to initial and booster vaccination, an overwhelming majority of both Delta and Omicron variant SARS-CoV-2 infections culture convert by day 6. Our work provides further support to the guidelines for strict masking beyond the initial 5-day isolation period for SARS-CoV-2 infections to help prevent transmission from the minority of cases who remain culture positive.

## Data Availability

All data produced in the present study are available upon reasonable request to the authors.

## Acknowledgement

The authors would like to acknowledge the Boston University students, faculty, and staff who have been impacted by the SARS-CoV-2 pandemic.

## Figure Legends

**Supplementary Figure 1:**
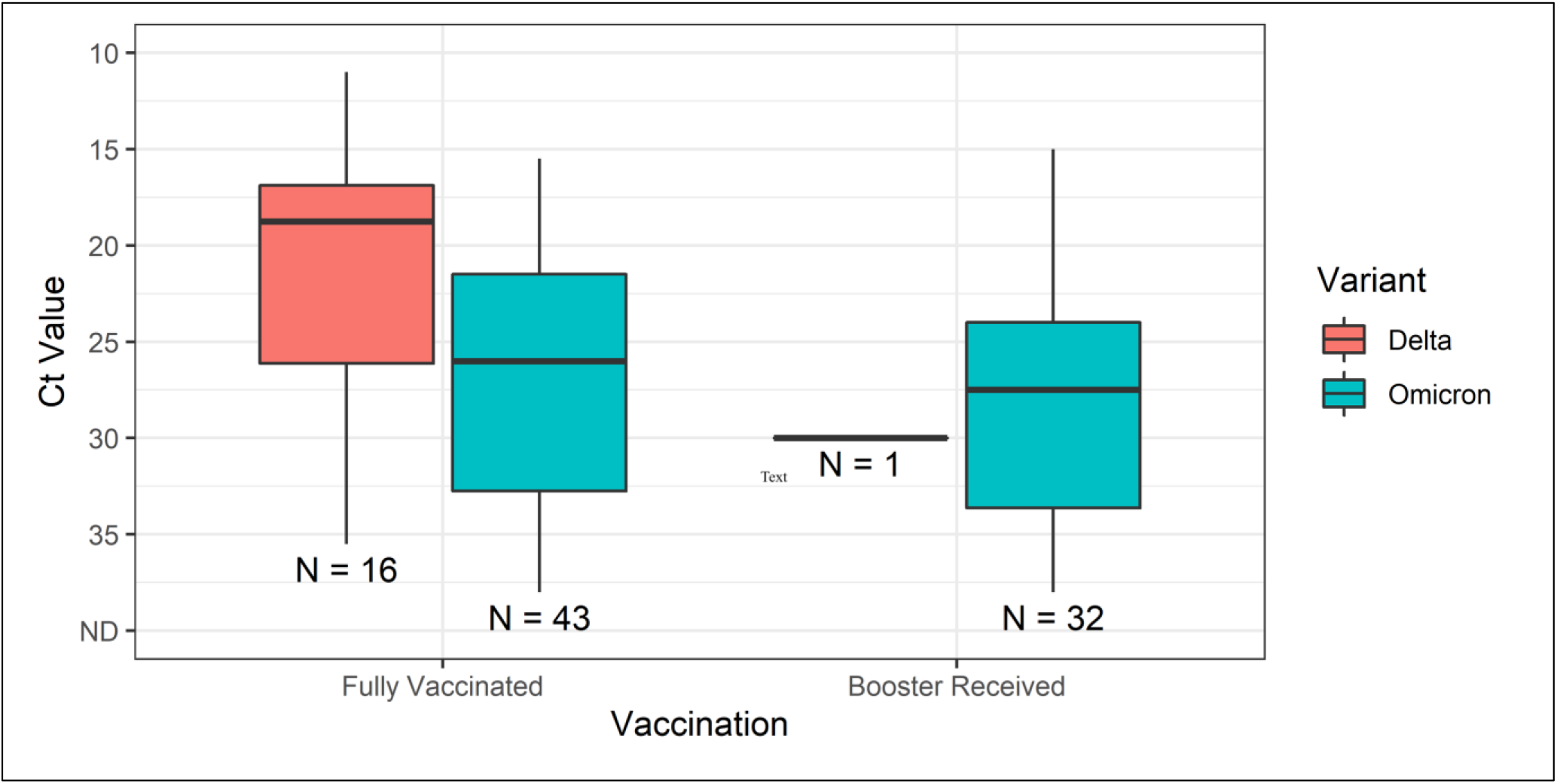
Box plots of polymerase chain reaction N1 cycle threshold by SARS-CoV-2 variant and vaccination status.

**Supplementary Figure 2:**
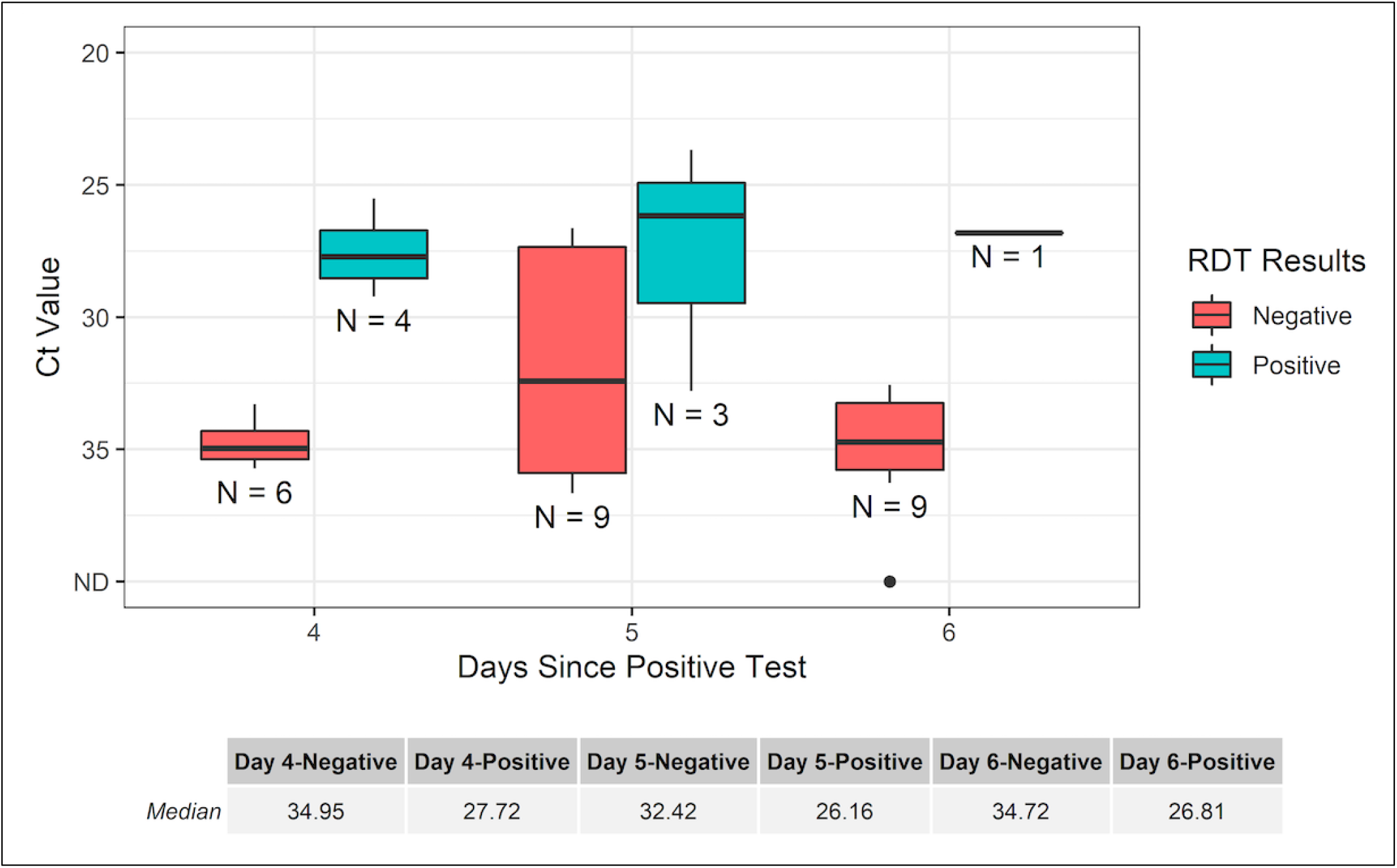
Box plots of polymerase chain reaction N1 cycle threshold by days from diagnosis and rapid diagnostic test result.

## References

1. CDC. Ending Isolation and Precautions for People with COVID-19: Interim Guidance. Centers for Disease Control and Prevention. Published January 14, 2022. Accessed March 17, 2022. https://www.cdc.gov/coronavirus/2019-ncov/hcp/duration-isolation.html

2. CDC Updates and Shortens Recommended Isolation and Quarantine Period for General Population. Centers for Disease Control and Prevention. Published December 29, 2021. Accessed March 17, 2022. https://www.cdc.gov/media/releases/2021/s1227-isolation-quarantine-guidance.html

3. Meyerowitz EA, Richterman A, Gandhi RT, Sax PE. Transmission of SARS-CoV-2: A Review of Viral, Host, and Environmental Factors. Ann Intern Med. 2021;174(1):69–79. doi:10.7326/M20-5008

4. Peeling RW, Heymann DL, Teo YY, Garcia PJ. Diagnostics for COVID-19: moving from pandemic response to control. Lancet. 2022;399(10326):757–768. doi:10.1016/S0140-6736(21)02346-1

5. Ettman CK, Abdalla SM, Cohen GH, Sampson L, Vivier PM, Galea S. Prevalence of Depression Symptoms in US Adults Before and During the COVID-19 Pandemic. JAMA Netw Open. 2020;3(9):e2019686. Published 2020 Sep 1. doi:10.1001/jamanetworkopen.2020.19686

6. Smith LE, Potts HWW, Amlôt R, Fear NT, Michie S, Rubin GJ. Adherence to the test, trace, and isolate system in the UK: results from 37 nationally representative surveys. BMJ. 2021;372:n608. Published 2021 Mar 31. doi:10.1136/bmj.n608

7. Nelson SB, Brenner IR, Homan E, et al. Evaluation of “Test to Return” after COVID-19 Diagnosis in a Massachusetts Public School District. Infectious Diseases (except HIV/AIDS); 2022. doi:10.1101/2022.02.11.22270843

8. Bays D, Whiteley T, Pindar M, et al. Mitigating Isolation: The Use of Rapid Antigen Testing to Reduce the Impact of Self-Isolation Periods. Public and Global Health; 2021. doi:10.1101/2021.12.23.21268326

9. Ito K, Piantham C, Nishiura H. Relative instantaneous reproduction number of Omicron SARS-CoV-2 variant with respect to the Delta variant in Denmark. Journal of Medical Virology. 2022;94(5):2265–2268. doi:10.1002/jmv.27560

10. Brandal LT, MacDonald E, Veneti L, et al. Outbreak caused by the SARS-CoV-2 Omicron variant in Norway, November to December 2021. Eurosurveillance. 2021;26(50). doi:10.2807/1560-7917.ES.2021.26.50.2101147

11. Petros BA, Turcinovic J, Welch NL, et al. Early Introduction and Rise of the Omicron SARS-CoV-2 Variant in Highly Vaccinated University Populations. Epidemiology; 2022. doi:10.1101/2022.01.27.22269787

12. Killingley B, Mann A, Kalinova M, et al. Safety, Tolerability and Viral Kinetics during SARS-CoV-2 Human Challenge. In Review; 2022. doi:10.21203/rs.3.rs-1121993/v1

13. Hamer DH, White LF, Jenkins HE, et al. Assessment of a COVID-19 Control Plan on an Urban University Campus During a Second Wave of the Pandemic. JAMA Netw Open. 2021;4(6):e2116425. doi:10.1001/jamanetworkopen.2021.16425

14. Bouton TC, Lodi S, Turcinovic J, et al. Coronavirus Disease 2019 Vaccine Impact on Rates of Severe Acute Respiratory Syndrome Coronavirus 2 Cases and Postvaccination Strain Sequences Among Health Care Workers at an Urban Academic Medical Center: A Prospective Cohort Study. Open Forum Infectious Diseases. 2021;8(10):ofab465. doi:10.1093/ofid/ofab465

15. Boucau J, Marino C, Regan J, et al. Duration of Viable Virus Shedding in SARS-CoV-2 Omicron Variant Infection. Infectious Diseases (except HIV/AIDS); 2022. doi:10.1101/2022.03.01.22271582

16. Hay JA, Kissler SM, Fauver JR, et al. Viral Dynamics and Duration of PCR Positivity of the SARS-CoV-2 Omicron Variant. Epidemiology; 2022. doi:10.1101/2022.01.13.22269257

17. Tai CG, Maragakis LL, Connolly S, et al. Booster Protection against Omicron Infection in a Highly Vaccinated Cohort. Infectious Diseases (except HIV/AIDS); 2022. doi:10.1101/2022.02.24.22271347

18. Hansen CH, Schelde AB, Moustsen-Helm IR, et al. Vaccine Effectiveness against SARS-CoV-2 Infection with the Omicron or Delta Variants Following a Two-Dose or Booster BNT162b2 or MRNA-1273 Vaccination Series: A Danish Cohort Study. Infectious Diseases (except HIV/AIDS); 2021. doi:10.1101/2021.12.20.21267966

19. Regan J, Flynn JP, Choudhary MC, et al. Detection of the Omicron Variant Virus with the Abbott BinaxNow SARS-CoV-2 Rapid Antigen Assay. Infectious Diseases (except HIV/AIDS); 2021. doi:10.1101/2021.12.22.21268219

20. Landon E, Bartlett AH, Marrs R, Guenette C, Weber SG, Mina MJ. High Rates of Rapid Antigen Test Positivity After 5 Days of Isolation for COVID-19. Infectious Diseases (except HIV/AIDS); 2022. doi:10.1101/2022.02.01.22269931

21. Lefferts B, Blake I, Bruden D, et al. Antigen Test Positivity After COVID-19 Isolation — Yukon-Kuskokwim Delta Region, Alaska, January–February 2022. MMWR Morb Mortal Wkly Rep. 2022;71(8):293–298. doi:10.15585/mmwr.mm7108a3

22. Cosimi LA, Kelly C, Esposito S, et al. Evaluation of the Role of Home Rapid Antigen Testing to Determine Isolation Period after Infection with SARS-CoV-2. Infectious Diseases (except HIV/AIDS); 2022. doi:10.1101/2022.03.03.22271766

